# Quantifying brain health in acute ischemic stroke through effective reserve

**DOI:** 10.1101/2024.03.22.24304714

**Authors:** Markus D. Schirmer, Kenda Alhadid, Robert W. Regenhardt, Natalia S. Rost

## Abstract

**Objective:** To quantify brain health using a measure of reserve that incorporates pre-existing pathology.

**Methods:** We analyzed two retrospective ischemic stroke cohorts (GASROS and SALVO) with neuroimaging and 90-day modified Rankin Scores (mRS) available. White matter hyperintensity (WMHv), brain, and intracranial volumes were automatically extracted, and brain parenchymal fraction (BPF) calculated. The latent variable effective reserve (eR) was modeled using age, WMHv, and BPF or brain volume in GASROS. Models were compared using Bayes Information Criterion (BIC). The best model’s eR estimates were categorized into quartiles and evaluated in SALVO.

**Results:** GASROS included 476 (median age: 65.8; 65.3% male) and SALVO included 43 (median age: 69.2; 62.8% male) patients. Inverse associations between eR and mRS was seen in both models, with brain volume outperforming BPF (path coefficients: -0.67, -0.48, respectively; p < 0.001; |ΔBIC| = 362). Quartile-based eR stratification in SALVO showed a similar inverse trend, with worse outcomes in the low reserve group (mRS≤2 - highest vs lowest quartile: 85/90% vs 59/45% for GASROS/SALVO).

**Conclusions:** Expanding the concept of eR, highlights its clinical translational potential. The strong link between higher eR and better outcomes underscores its value as a protective brain health metric.

## Introduction

Stroke is a leading cause of long-term disability worldwide making the prevention of related physical and cognitive impairment essential.^1^ Comprehensive outcome modeling may lead to effective prevention strategies for adverse outcomes, enriching quality of life and reducing economic burden.^2^ However, recovery mechanisms are complex, and current models are limited.^3^

With brain health becoming a significant action goal, reserve concepts can help explain outcome differences.^4,5^ While they are established in fields such as neurodegeneration, their adaptation to stroke populations is new.^6,7^ In an initial study,^8^ we expanded the idea of structural reserve in stroke, introducing *effective reserve* (eR) as a latent variable which accounts for pre-existing pathology. However, clinical neuroimaging challenges, such as low image quality and acquisition variability, remain when quantifying existing neuropathology. Recently, we developed automated approaches to quantify white matter hyperintensity (WMH) and brain volume in these settings,^9-11^ which enable rapid quantification of these important biomarkers at the bedside.

This study sought to refine our model of eR, which relied on measures of intracranial volume (ICV) and systolic blood pressure at the time of admission, by incorporating brain volume and WMH volume instead, and to demonstrate that the protective mechanism of effective reserve offsets the negative effect of the stroke lesion. We aim to demonstrate that measures of eR can be easily derived from standard-of-care neuroimaging, allowing a quartile-based prognostication that can be widely implemented as an accessible tool across stroke centers.

## Methods

### Standard protocol approvals, registration, and patient consent

The study cohorts received approval from the local Institutional Review Board, and informed written consent was obtained from all patients or their surrogates, in line with the Declaration of Helsinki.

### Study design and neuroimaging data

Patients from the GASROS^12^ (2003-2011) and SALVO^13^ (2014-2019) cohorts with acute ischemic stroke confirmed on diffusion-weighted imaging (DWI) within 48 hours, and with available T2 fluid-attenuated inversion recovery (T2-FLAIR), were included. Demographics and medical history were recorded on admission. Functional outcomes were assessed at 3 months using the modified Rankin Scale (mRS), based on interviews with patients/caregivers or review of clinical evaluations.

GASROS MRI data included DWI (single-shot echo-planar imaging; 1-5 B0 volumes, 6-30 diffusion directions with b=1000 s/mm^2^, 1-3 averaged volumes) and acute infarct volume was manually quantified. Both GASROS and SALVO patients had T2 FLAIR imaging (TR 5000ms, TE 62-116ms, TI 2200ms, FOV 220-240mm), as part of the standard protocol. Automated estimates of WMH,^9^ along with ICV and brain volume (combined white and gray matter),^11^ were determined using image analysis pipelines developed for acute stroke.

### Statistical analysis and model description

Each segmentation underwent manual quality control by visual inspection. WMH, brain, and ICV were calculated by multiplying voxel number by voxel size. Brain parenchymal fraction (BPF) was calculated as the ratio of brain volume to ICV in GASROS and logit-transformed. WMH and lesion loads were given as the ratio of WMH and lesion volume to brain volume and logit-transformed. Brain volume was reported in dm^3^ and age in decades.

eR was modeled using latent variable analyses,^8^ given by

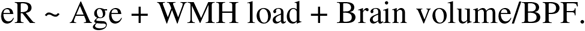

The outcome model also included stroke lesion load, sex, hypertension, diabetes mellitus, and smoking as predictors. Parameters were estimated using the R package LAVAAN,^14^ and models were compared using the Bayes Information Criterion (BIC) using the GASROS cohort.

eR was then inferred in both GASROS and SALVO using the path-coefficients of the model with the lowest BIC. Cohort outcome distributions were stratified by quartiles and compared using the Kolmogorov-Smirnov test. All analyses were performed in R,^15^ with significance set at p<0.05.

### Data availability statement

Data, methods, and materials will be made available to any researcher for the purpose of reproducing the results, subject to approval by the local Institutional Review Board.

## Results

Table 1 outlines the cohort characteristics. In the GASROS cohort (n = 476), the median age was 65.8 years (IQR: 55.3-76.3), 65.3% were male, and 69.7% had hypertension. In the SALVO cohort (n = 43), the median age was 69.2 years (IQR: 61.2-76.3), 62.8% were male, and 67.4% had hypertension. Both cohorts had similar median 90-day mRS.

**Table 1.**
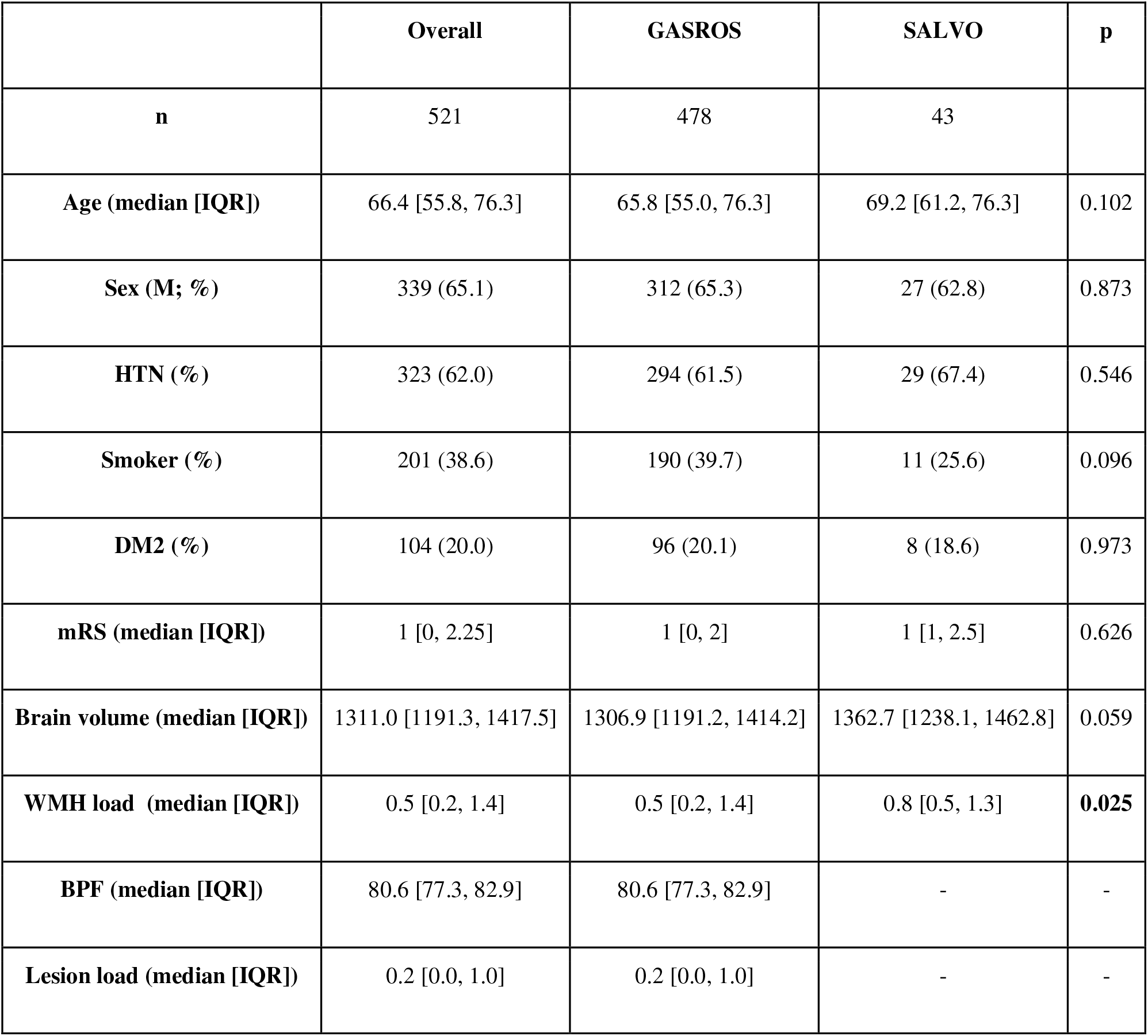
Characteristics of the cohort utilized in this study. (IQR: interquartile range; HTN: hypertension; DM2: Diabetes Mellitus Type 2; BPF: brain parenchymal fraction)

|  | Overall | GASROS | SALVO | p |
| --- | --- | --- | --- | --- |
| **n** | 521 | 478 | 43 |  |
| **Age (median [IQR])** | 66.4 [55.8, 76.3] | 65.8 [55.0, 76.3] | 69.2 [61.2, 76.3] | 0.102 |
| **Sex (M; %)** | 339 (65.1) | 312 (65.3) | 27 (62.8) | 0.873 |
| **HTN (%)** | 323 (62.0) | 294 (61.5) | 29 (67.4) | 0.546 |
| **Smoker (%)** | 201 (38.6) | 190 (39.7) | 11 (25.6) | 0.096 |
| **DM2 (%)** | 104 (20.0) | 96 (20.1) | 8 (18.6) | 0.973 |
| **mRS (median [IQR])** | 1 [0, 2.25] | 1 [0, 2] | 1 [1, 2.5] | 0.626 |
| **Brain volume (median [IQR])** | 1311.0 [1191.3, 1417.5] | 1306.9 [1191.2, 1414.2] | 1362.7 [1238.1, 1462.8] | 0.059 |
| **WMH load (median [IQR])** | 0.5 [0.2, 1.4] | 0.5 [0.2, 1.4] | 0.8 [0.5, 1.3] | **0.025** |
| **BPF (median [IQR])** | 80.6 [77.3, 82.9] | 80.6 [77.3, 82.9] | - | - |
| **Lesion load (median [IQR])** | 0.2 [0.0, 1.0] | 0.2 [0.0, 1.0] | - | - |

Structural equation models, including their path coefficients, are shown in Figure A1 and Table A1. Model comparison using BIC resulted in |ΔBIC| = 362 (brain volume model BIC=4462; BPF model BIC=4824) with a path coefficient of -0.67 between eR and mRS. Model parameters of the brain volume model further suggest that age and WMH load negatively affect eR (path coefficients -0.85 and -0.51, respectively; p<0.001), whereas higher brain volume leads to an increase in eR (path coefficient 0.1; p<0.001). The model showed that eR has a 2.8 times larger, opposite effect, compared to lesion load (path coefficient: 0.24).

eR values were calculated in both GASROS and SALVO and quartiles determined in both cohorts. The outcome distributions per quartile are shown in Figure 1, demonstrating patients in higher eR quartiles generally had better 90-day mRS (mRS≤2 - highest quartile: 85/90%; lowest quartile: 59/45% for GASROS/SALVO). The Kolmogorov-Smirnov test did not identify a significant difference in outcome distributions within quartiles between cohorts.

**Figure 1.**
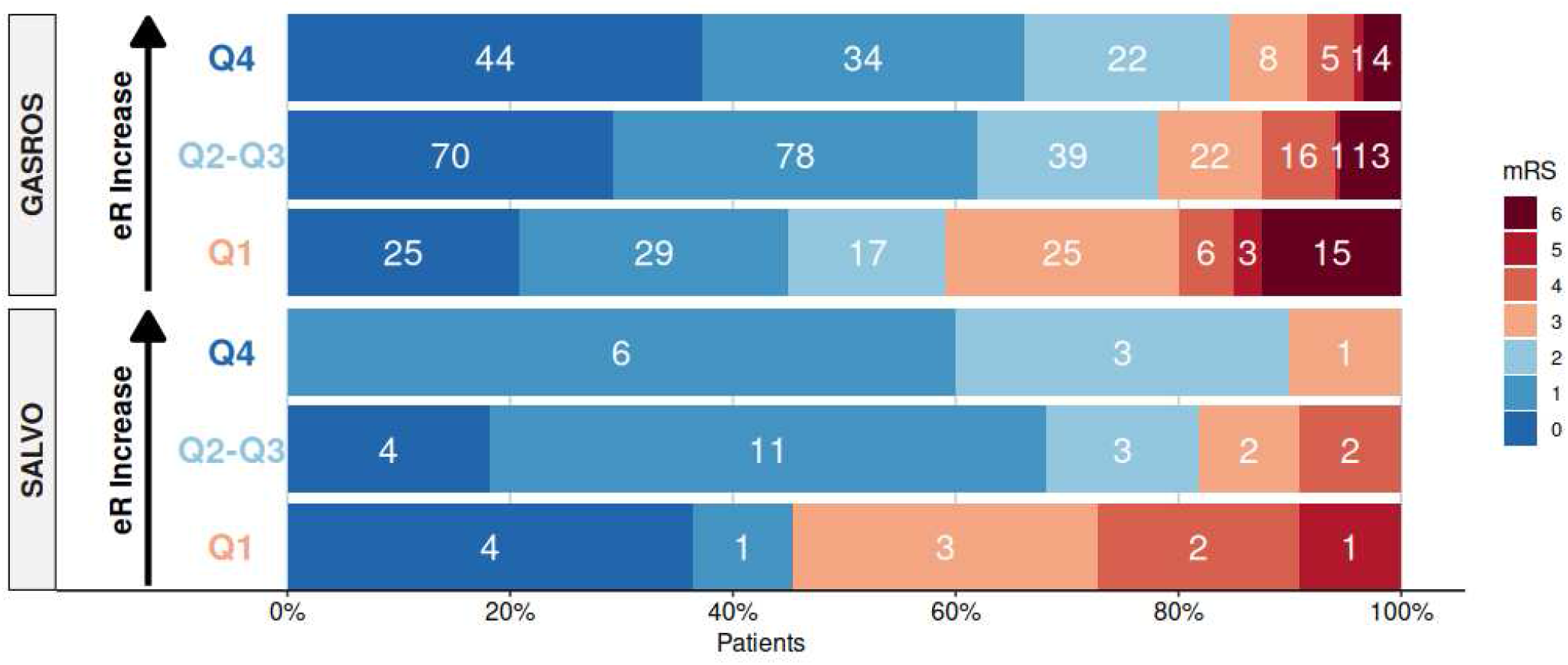
Quartile distribution of modified Rankin Scale (mRS) in GASROS and SALVO. Good outcome (mRS≤2) in shown shades of blue. Quartiles in each cohort based on eR, calculated using the path coefficients of the structural equation model in GASROS, represent high (Q4), intermediate (Q2-Q3), and low (Q1) brain health. White numbers reflect the number of patients per mRS category, with higher quartiles generally correlating with better outcomes.

## Discussion

By quantifying the brain’s capacity to withstand insults, eR is a surrogate measure of brain health. Here, we extended this concept incorporating information related to brain health and linking eR to long-term outcome.

Using deep-learning neuroimage analyses,^9,11^ we quantified ICV, brain, and WMH volumes from clinically available MRI. Higher eR estimates led to better outcomes, where the brain volume model substantially outperformed the BPF model (|ΔBIC| = 362>10). This result aligns with recent research showing brain volume outperforms BPF in multivariable linear regression models.^11^ We note that eR has 2.8 and 1.9 times larger effect on mRS in amplitude, but opposite in direction, compared to lesion load in the brain volume and BPF models. Using a quartile-based categorization of eR, patients in higher eR quartiles consistently showed better functional outcomes in both cohorts. This result further underscores the potential of eR for characterizing a significant protective mechanism and its importance for outcome prognostication.

There are some limitations to the current study. mRS tends to over-emphasize motor function, lacking comprehensive cognitive and patient centric outcome information. Additionally, mRS obtained through interviews with patients and/or caregivers can introduce self-report biases. However, it remains a standard outcome measure after stroke, supporting the translational potential of our results. Moreover, stroke treatment information was unavailable, but likely consisted of thrombolytic therapy. Future studies with other measures of outcomes and treatment information are needed to fully investigate the potential of eR as a pivotal marker of brain health.

In this study, we significantly extended the concept of eR and demonstrated its utility as an important biomarker for stroke outcome. Utilizing routinely acquired clinical neuroimaging data supports the immediate translational potential of the results, allowing us to create a quantitative marker of brain health and evaluate its relationship to post-stroke outcomes at the time of presentation. The association of higher eR with better post-stroke outcome highlights its potential to be used as a quantitative, easily extractable measure of brain health, and as an indicator of the brain’s protective mechanisms against acute ischemic injury.

## Data Availability

The authors agree to make the data, methods used in the analysis, and materials used to conduct the research available to any researcher for the express purpose of reproducing the results and with the explicit permission for data sharing by the local institutional review board.

## Appendix

**Figure A1.**
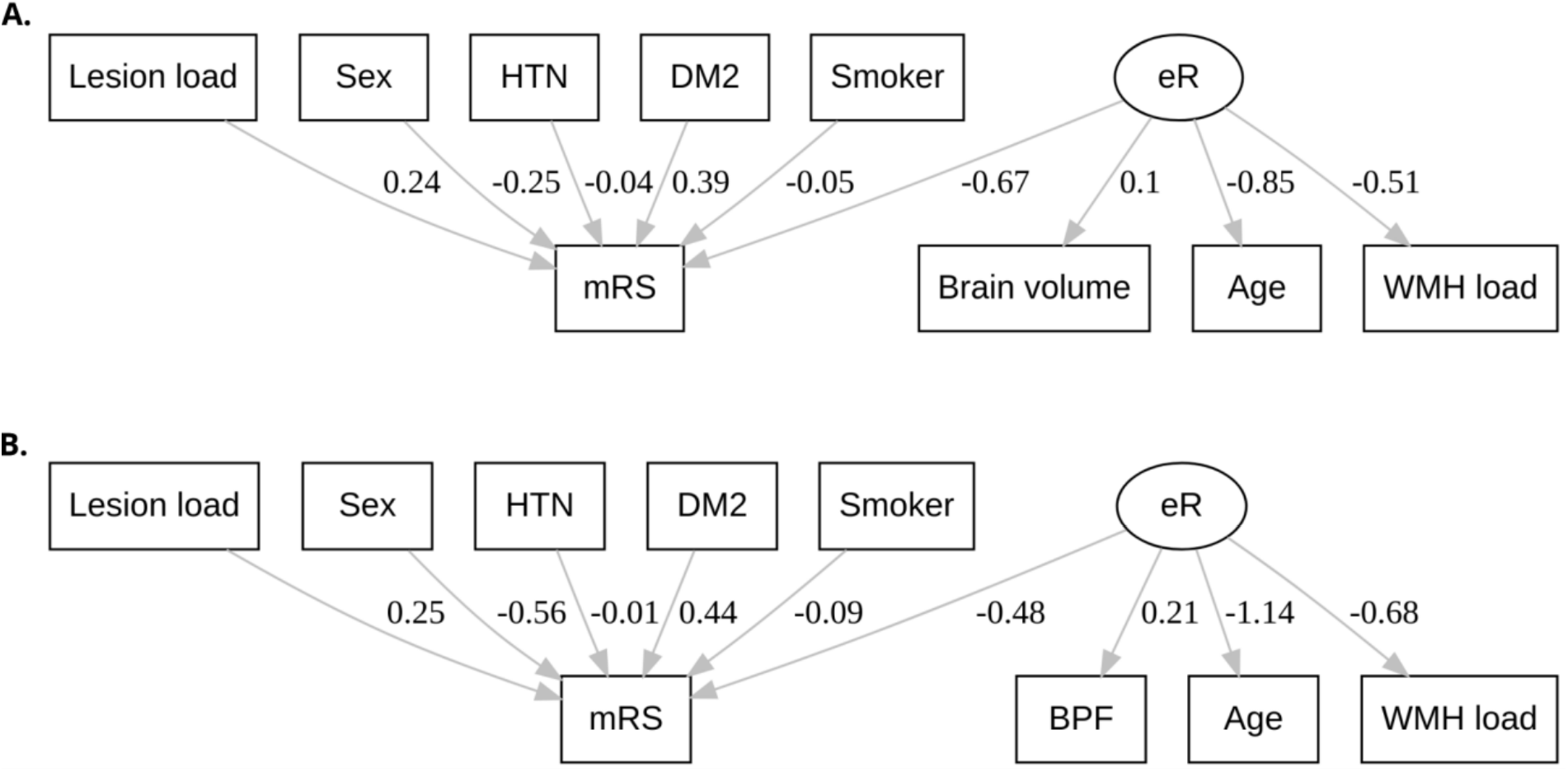
Structural equation models with estimated associations using path analysis. Model parameters of the brain volume model (Figure AlA) suggest that age and white matter hyperintensity (WMH) load negatively affect effective reserve (eR; path coefficients -0.85 and - 0.51, respectively; p<0.001), whereas higher brain volume leads to an increase in eR (path coefficient 0.1; p<0.001). The brain parenchymal fraction (BPF) model (Figure AlB) shows the same trends, with age and WMH load reducing eR (path coefficients -1.14 and -0.68, respectively; p<0.001), and higher BPF, i.e., less brain atrophy, leading to a higher effective reserve. All path coefficients had a p-value of p<0.001 with the brain volume model (A; BIC=4462) outperforming the BPF model (B; BIC=4824). HTN: hypertension; DM2: Diabetes Mellitus Type 2;

**Table A1.**
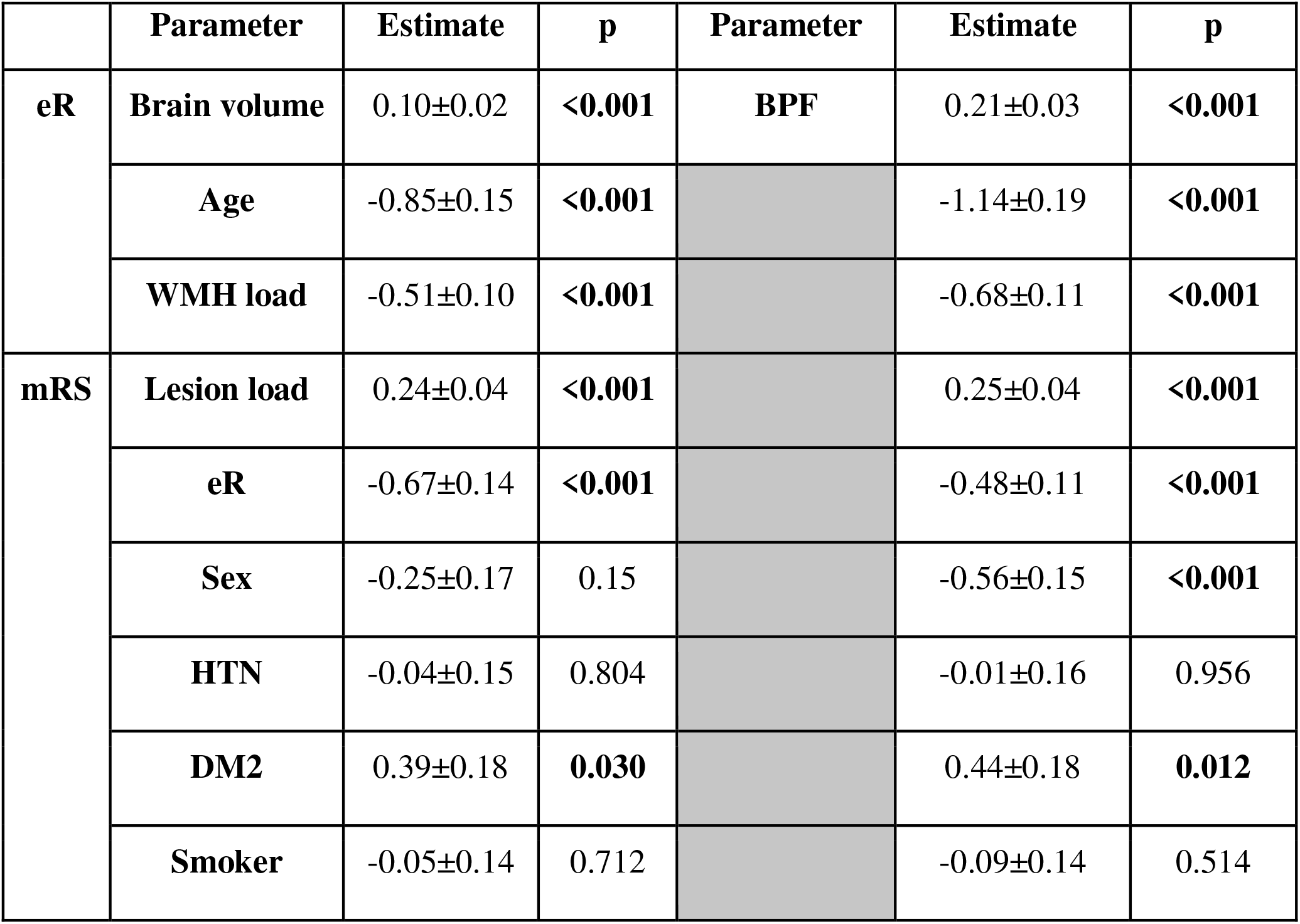
Path-coefficients corresponding to Figure A1. (WMH: white matter hyperintensity; eR: effective reserve; HTN: hypertension; DM2: Diabetes Mellitus Type 2; BPF: brain parenchymal fraction)

|  | Parameter | Estimate | p | Parameter | Estimate | p |
| --- | --- | --- | --- | --- | --- | --- |
| **eR** | **Brain volume** | 0.10±0.02 | **<0.001** | **BPF** | 0.21±0.03 | **<0.001** |
|  | **Age** | -0.85±0.15 | **<0.001** |  | -1.14±0.19 | **<0.001** |
|  | **WMH load** | -0.51±0.10 | **<0.001** |  | -0.68±0.11 | **<0.001** |
| **mRS** | **Lesion load** | 0.24±0.04 | **<0.001** |  | 0.25±0.04 | **<0.001** |
|  | **eR** | -0.67±0.14 | **<0.001** |  | -0.48±0.11 | **<0.001** |
|  | **Sex** | -0.25±0.17 | 0.15 |  | -0.56±0.15 | **<0.001** |
|  | **HTN** | -0.04±0.15 | 0.804 |  | -0.01±0.16 | 0.956 |
|  | **DM2** | 0.39±0.18 | **0.030** |  | 0.44±0.18 | **0.012** |
|  | **Smoker** | -0.05±0.14 | 0.712 |  | -0.09±0.14 | 0.514 |

## References

1. Jacquin A, Binquet C, Rouaud O, et al. Post-stroke cognitive impairment: high prevalence and determining factors in a cohort of mild stroke. J Alzheimers Dis. IOS Press; 2014;40:1029–1038.

2. World Health Organization. Neurological disorders: public health challenges. WHO; 2006.

3. Brugnara G, Neuberger U, Mahmutoglu MA, et al. Multimodal predictive modeling of endovascular treatment outcome for acute ischemic stroke using machine-learning. Stroke. Am Heart Assoc; 2020;51:3541–3551.

4. Mortimer JA, Borenstein AR, Gosche KM, Snowdon DA. Very early detection of Alzheimer neuropathology and the role of brain reserve in modifying its clinical expression. J Geriatr Psychiatry Neurol. Sage Publications Sage CA: Thousand Oaks, CA; 2005;18:218–223.

5. Valenzuela MJ, Sachdev P. Brain reserve and dementia: a systematic review. Psychol Med. 2006;36:441–454.

6. Rosenich E, Hordacre B, Paquet C, Koblar SA, Hillier SL. Cognitive reserve as an emerging concept in stroke recovery. Neurorehabil Neural Repair. SAGE Publications Sage CA: Los Angeles, CA; 2020;34:187–199.

7. Shin M, Sohn MK, Lee J, et al. Effect of cognitive reserve on risk of cognitive impairment and recovery after stroke: the KOSCO study. Stroke. Am Heart Assoc; 2020;51:99–107.

8. Schirmer MD, Etherton MR, Dalca AV, et al. Effective reserve: a latent variable to improve outcome prediction in stroke. J Stroke Cerebrovasc Dis. 2019;28:63–69.

9. Schirmer MD, Dalca AV, Sridharan R, et al. White matter hyperintensity quantification in large-scale clinical acute ischemic stroke cohorts-The MRI-GENIE study. NeuroImage Clin. Elsevier; 2019;23:101884.

10. Oliveira LC, Bonkhoff AK, Regenhardt RW, et al. Neuroimaging markers of patient-reported outcome measures in acute ischemic stroke. MedRxiv Prepr. Serv. Health Sci. 2023. p. 2023.12.27.23299829.

11. Alhadid K, Regenhardt RW, Rost NS, Schirmer MD. Brain volume is a better biomarker of outcomes in ischemic stroke compared to brain atrophy. arXiv; 2024. p. 2403.12788.

12. Zhang CR, Cloonan L, Fitzpatrick KM, et al. Determinants of white matter hyperintensity burden differ at the extremes of ages of ischemic stroke onset. J Stroke Cerebrovasc Dis. Elsevier; 2015;24:649–654.

13. Ktena SI, Schirmer MD, Etherton MR, et al. Brain connectivity measures improve modeling of functional outcome after acute ischemic stroke. Stroke. Am Heart Assoc; 2019;50:2761–2767.

14. Rosseel Y. lavaan: An R package for structural equation modeling. J Stat Softw. 2012;48:1–36.

15. R Core Team. R: A language and environment for statistical computing. Vienna, Austria; 2024.

